# Does the Data Tell the True Story? A Modelling Study of Early COVID-19 Pandemic Suppression and Mitigation Strategies in Ghana

**DOI:** 10.1101/2021.05.02.21256473

**Authors:** Nana Kena Frempong, Theophilus Acheampong, Ofosuhene O. Apenteng, Emmanuel Nakua, John H. Amuasi

## Abstract

This paper uses publicly available data and various statistical models to estimate the basic reproduction number (*R*_*0*_) and other disease parameters for the early COVID-19 pandemic outbreak in Ghana. We also test the effectiveness of government imposition of public health measures to reduce the risk of transmission and impact of the pandemic, especially in the early phase. R_0_ is estimated from the statistical model as 3.21 using a 0.147 estimated growth rate [95% C.I.: 0.137-0.157] and a 15-day time to recovery after COVID-19 infection. This estimate of the initial R_0_ is consistent with others reported in the literature from other parts of Africa, China and Europe. Our results also indicate that COVID-19 transmission reduced consistently in Ghana after the imposition of public health interventions - such as border restrictions, intra-city movement, quarantine and isolation - during the first phase of the pandemic from March to May 2020. However, the time-dependent reproduction number (R_t_) beyond mid-May 2020 does not represent the true situation given there was not a consistent testing regime in place. This is also confirmed by our Jack-knife bootstrap estimates which show that the positivity rate over-estimates the true incidence rate from mid-May 2020. Given concerns about virus mutations, delays in vaccination and a possible new wave of the pandemic, there is a need for systematic testing of a representative sample of the population to monitor the reproduction number. There is also an urgent need to increase the availability of testing for the general population to enable early detection, isolation and treatment of infected individuals to reduce progression to severe disease and mortality.

## 1 Introduction

The Coronavirus disease (COVID-19), a novel infectious disease caused by the Severe Acute Respiratory Syndrome coronavirus 2 (SARS-CoV-2) was declared a global pandemic by the World Health Organization (WHO) on March 11, 2020 (Cucinotta and Vanelli, 2020; WHO, 2020a). The first reported case was in December 2019 in Wuhan, the capital of China’s Hubei province, from where it quickly spread globally, reaching Sub-Saharan Africa (SSA) including Ghana (Leung et al, 2020; Cowling et al., 2020). The Africa Centre for Disease Control (AfCDC) on April 7, 2021 indicated that the continent had recorded 3.03 million cases of the virus with 2.75 million recoveries and 78,005 deaths. This represented 2.28% of all global cases (133.07 million), 2.57% of global recoveries (107.27 million), and 2.7% of total global deaths (2.89 million) as of April 07, 2021.

Ghana reported its first case of COVID-19 on March 12, 2020. Since then, the Government of Ghana had reported 91,009 total confirmed cases with 88,810 recoveries and 752 deaths as of April 07, 2021 (Ghana Health Service, 2021; Africa CDC, 2021). In response to the outbreak, the Ghanaian government, like other sovereign nation states, imposed public health measures such as restrictions on movement, including lockdowns to contain the spread of the virus. The country closed land, air and sea borders in mid-March 2021. In addition, all educational institutions, including universities, high schools, and primary schools (both public and private) were closed. The authorities also imposed bans on public gatherings. The government subsequently on April 20, 2020, lifted a three-week lockdown on the movement of persons within some parts of the Greater Accra, Tema, Kasoa and Greater Kumasi metropolis.

Preliminary evidence of the impact of these restrictions on the mobility patterns of populations for the period between February 17 and May 03, 2021 using anonymised and aggregated data from Ghana’s Mobile Network Operators (MNOs), indicated that person movements decreased up to 60% relative to the baseline value (Ghana Statistical Service, 2020). For example, in the Greater Accra Region, where about 70% of the total cases were located at the time, person movements decreased by 20-30% during the period when initial restrictions were put in place as compared to the baseline value (Ghana Statistical Service, 2020). This further decreased by 50-60% during the 3-week lockdown period, relative to the baseline level. A similar trend was observed for the Kumasi Metropolis which had 15% of the total cases at that time (Ghana Statistical Service, 2020).

There is a natural expectation that the lockdown and other control measures would have had some positive effect in restricting movement and physical interaction and thereby possibly slowing the spread of the virus. However, there is a need to fully explore using data, what the direction of impact and magnitude of these interventions truly was, especially in the early stages of the COVID-19 pandemic in Ghana. The emphasis on the early period of the pandemic covering March to July 2020 is particularly important because within this period, Ghana was among the few countries in Africa actively testing suspected cases and their contacts via a Tracing, Testing, and Treatment (‘3Ts’) programme and publishing the positivity rate, (Sibiri et al., 2020; Zhang et al, 2020). For example, as at April 25, 2020, Ghana had the highest COVID-19 test per capita in SSA with a rolling 7-day average of 0.18 daily new COVID-19 tests per 1,000 people compared to South Africa’s 0.13 and Senegal’s 0.03 (OurWorldinData, 2021). Some of the response strategies, such as tracing and testing of contacts, were subsequently scaled back, largely due to financing constraints and the continued spread. Nevertheless, data from this early period (March to July 2020) is relatively more accurate and reliable as reflecting the epidemiology of the disease in Ghana than that collected much later, because in this early period, testing was centralized and compliance with timely reporting was high.

To formulate a valid and reliable estimate of transmissibility and spread of the COVID-19 outbreak in Ghana, we have utilised statistical modelling, which facilitates understanding of the mathematical determination of how the disease spreads. In epidemiological studies, several mathematical and statistical models have been proposed to understand the transmission dynamics of infectious diseases (Kramer et al., 2010; Diekman & Heesterbeek; 2000, Brauer & Castillo-Chavez, 2012; Gilbert et al., 2020; Diop et al., 2020).

Modelling an infectious disease such as COVID-19 in an African country with reliable data is critical to understanding the transmission dynamics so this knowledge can be applied to planning and decision-making at various levels.

To assess the intensity of an outbreak, transmissibility can be quantified by the basic reproduction number (R_0_), that is, the average number of secondary infections generated by a single primary infective in a completely susceptible population (Diekmann et al., 1990). The importance of being able to estimate the basic reproduction number (R_0_) becomes more apparent when an emerging infectious disease strikes a population and there is need for a tailored intervention. Monitoring the basic reproduction number provides insight into how the spread of COVID-19 is impacted by various events/interventions and is critical for decision-making in introducing or modifying (including lifting) social-distancing measures and their health and socio-economic implications (Ma et al., 2014). R_0_ is, therefore, the fulcrum of any predictive modelling regarding Ghana’s COVID-19 epidemic curve, including any effort to estimate the impact of expected events which modify human movement and contact and most importantly, any mitigation strategies. The R_0_ is therefore one of governments’ key metrics for determining how well otherwise disruptive disease-spread mitigation strategies like lockdowns (of various forms) and other policies worked and might work in the future. We can also use the monitoring of the R_0_ to determine if these polices are worth the socio-economic costs they impose, especially in a largely informal economy such as Ghana’s (Danquah & Schotte, 2020). R_0_ is a dynamic value that can vary geographically and temporally (Ridenhour et al., 2018; Ng & Wen, 2019.). It can be different from one day to another and from one place to another for the same disease, and its increase or decrease is determined by local conditions and human behaviour (Delamater et al., 2019; Dharmaratne et al., 2020).

Given the foregoing, the aim of this paper is to estimate the basic reproduction number (R_0_) and other disease parameters for the early evolution of the COVID-19 pandemic in Ghana. We also use our estimations to explain the epidemiology of the disease in Ghana and explore the effectiveness of the government’s imposition of public health measures to suppress and mitigate the further spread of the pandemic in the country from March to July 2020. We develop a statistical model for the initial COVID-19 outbreak using various parametric growth models for the daily new cases in Ghana based on publicly available data. Additionally, we estimate the evolution of the reproduction number before a local maximum incidence of the COVID-19 pandemic in the country. Two methods are used for the estimation of the evolving R_0_, namely Sequential Bayesian (SB) and Time-Dependent (TD). The Jack-knife estimation technique is also applied to the data to examine any inconsistencies in testing rates within the period under review.

## 2 Methods

### 2.1 Data sources

Data for the COVID-19 pandemic in Ghana was obtained from the Ghana Health Service, which is an agency operating under the Ministry of Health, Government of Ghana. The first 135 days after first case was reported (March 12, 2020 to July 24, 2020) was used for the analysis as the first phase of the epidemic in Ghana. Figure 1 shows COVID-19 indicators for Ghana from March to December 2020.

**Figure 1:**
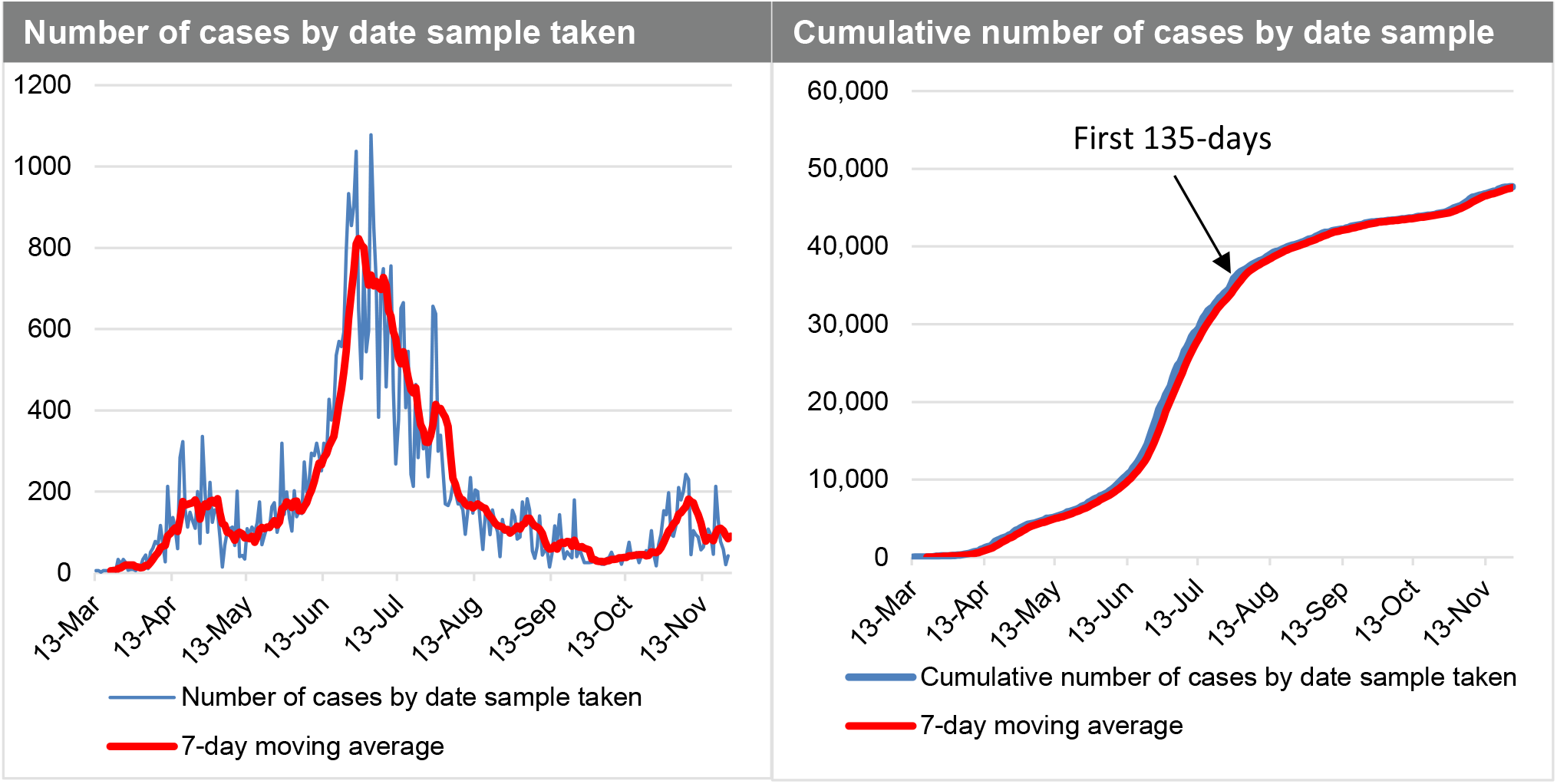
Ghana COVID-19 daily cases (Mar-Dec 2020) Data source: Ghana Health Service (2020)

### 2.2 Statistical model for the initial COVID-19 outbreak in Ghana

We focus on the initial stage of the epidemic using daily incidence data from the date of first reported case in the country. The weekly reported cases used was to attenuate any potential bias introduced by reporting delays. We let *Y(t)* be the number of COVID-19 cases at time *t*. We assume that

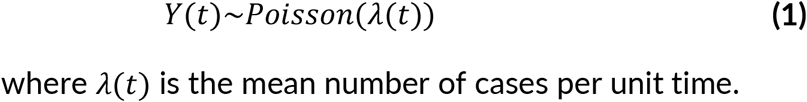

We assume that in the initial stages of the epidemic, the daily incidence cases follow some growth models. The following are the models considered to estimate the growth rate of the initial stage of the epidemic.

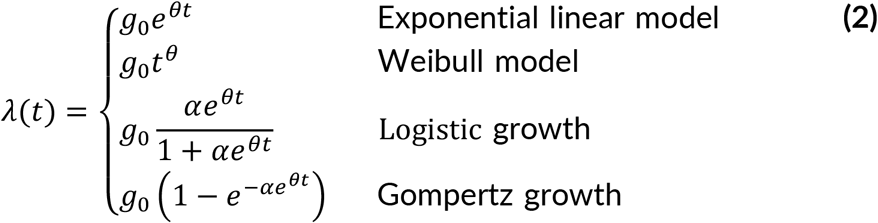

We note that at *t* > 0, the initial number of cases *g*_0_ is a constant and used as a parameter in the model. The parameter represents the growth rate of the epidemic. An estimate of the growth rate would help to estimate the basic reproduction number for proposed mathematical model of COVID-19 (Ma, 2020).

#### 2.2.1 Estimating initial R0 under a Generalised Linear Model (GLM) framework

The number of COVID-19 cases at time *t* modelled falls under the GLM framework (McCullagh and Nelder, 1989) with Poisson distribution for the response and a log link function. The exponential linear and Weibull models in Equation (2) would be formulated under the GLM while the log-logistic and Gompertz growth models are formulated under non-linear models. Using the log link, we transform the mean function of both exponential and Weibull models to a linear structure. We assume the number of cases *y*(*t*_*i*_) at time *t*_*i*_ is independently Poisson distributed with mean 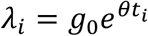.

The log-likelihood function is given as:

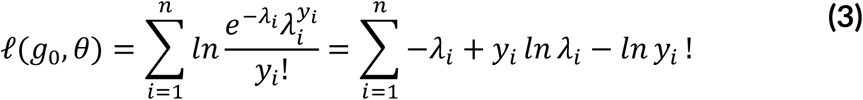

Maximizing the log-likelihood ignoring terms independent on the parameters and substituting *λ*_*i*_ in (3).

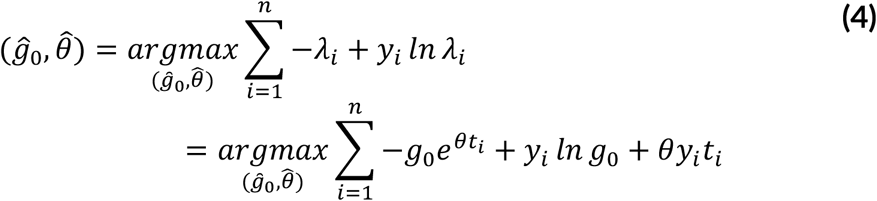

Similarly, for 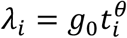 the log-likelihood is given as

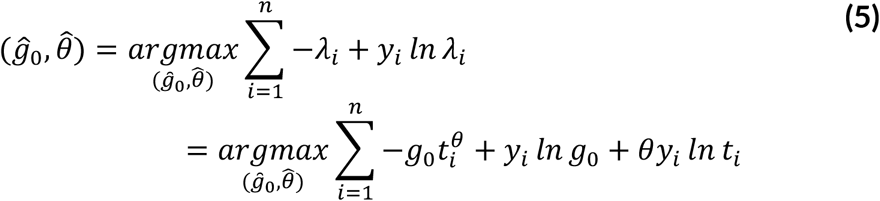

for 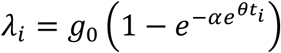, the log-likelihood is given as

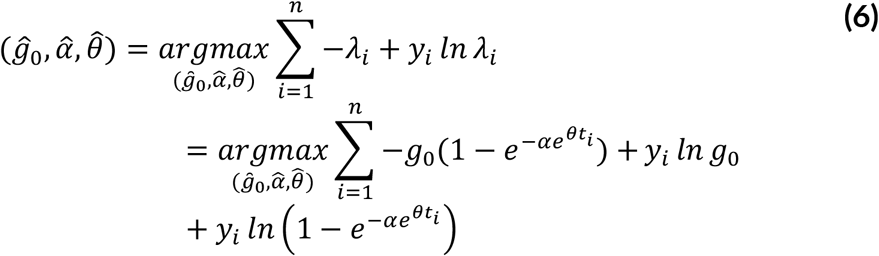

For 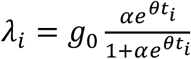, the log-likelihood is given as

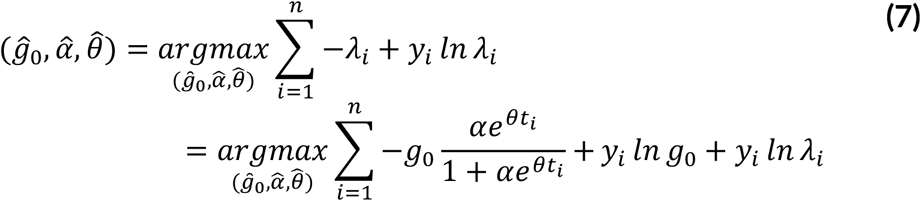

This maximisation problem is solved numerically using the Newton-Raphson’s algorithm.

The basic reproduction number *R*_0_is closely related to the initial growth rate *θ* under the assumption of exponentially distributed latent and infectious periods using the formula (Feng, 2007):

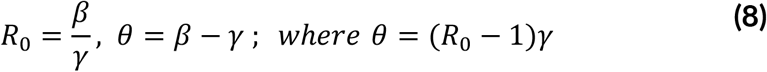

where *β,γ* are the transmission and recovery rates respectively

#### 2.2.3 Estimating R_0_ over time before a local maximum incidence of COVID-19

Two methods were considered for the estimation of *R*_*0*,_namely sequential Bayesian (SB) and Time-Dependent (TD). These two methods rely on the generation time, which is the time lag between infection in a primary case and transmission to a secondary case. The generation time distribution is obtained from the time lag between all infective/infector pairs, as it cannot be observed directly from the data. So, we often substitute with the serial interval distribution that measures time between symptom onset. The implementation was done in the R software using the *R*_*0*_ package implemented by Obadia et al. (2012). These methods rely on the assumption that the number of secondary cases caused by an index case is Poisson distributed. Given observation of (*N*_0_, *N*_1_, …, *N*_*T*_) incident cases over consecutive time units, and a generation time distribution w, *R*_*0*_ is estimated by these two methods:

##### Sequential Bayesian method (SB)

This method relies on an approximation to the SIR model, whereby incidence at time *t* + 1, (*t* + 1) is approximately Poisson distributed with mean

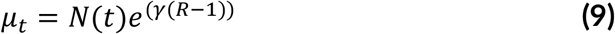

Bayesian framework starts with a non-informative prior on the distribution of the reproduction number *R*_*0*_.

The distribution is updated as new data is observed, using

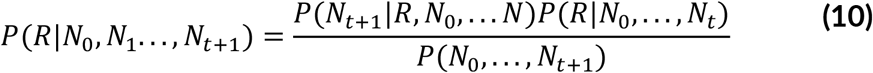

##### Estimation of time-dependent reproduction numbers (TD)

The time-dependant method proposed by Wallinga & Teunis (2004), computes reproduction numbers by averaging over all transmission networks compatible with observations. The probability *p*_*ij*_ that case *i* with onset at time *t*_*i*_ was infected by case *j* with onset at time *t*_j_ is calculated as

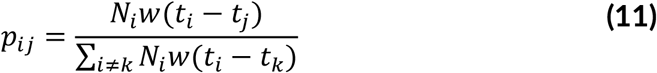

The effective number for case *j* is therefore *R*_*j*_ = Σ _*j*_ *p*_*ij*_ and averaged as 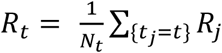 over all cases with same date of onset.

#### 2.2.3 Estimating the impact of easing restrictions on infection rates

Fokianos and Fried (2010, 2012) proposed intervention time series, and models interventions affecting the location by including a deterministic covariate and a decay rate. Following Fokianos and Fried (2010, 2012), we let *Y*_*t*_ be the number of COVID-19 cases at time *t*. We assume that *Y*_*t*_ *∽ NegBin* (*λ*_*t*_ *ϕ*), where *λ*(*t*) is the mean number of cases per unit time and *ϕ* the dispersion parameter. In general. the linear predictor of a model with *S* types of interventions according to parameters *δ*_1_,…, *δ*_*S*_ occurring at time points τ_1_,…, τ_*S*_ where, *w*_*m*_, m = 1,…, *S* are intervention sizes is given as follows:

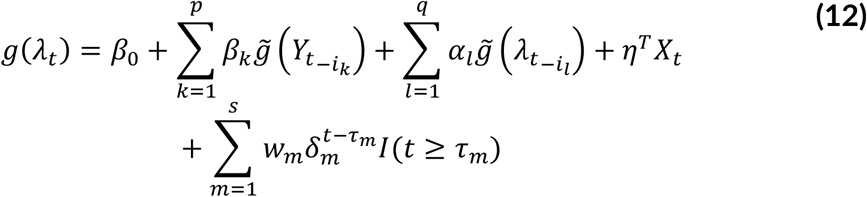

The maximum likelihood estimation (MLE) is used to obtain the parameter estimates.

By identifying interventions and estimating their effect size, we allow for the identification of structural changes which can be assessed using valid statistical inferences. That is, did the said intervention have an effect on a time series during the respective period(s) in question. This is a test under the null hypothesis (H_0_) of no intervention, against the alternative hypothesis (H_1_) of some intervention at a known time τ. If the policy interventions actually worked, then we should see a statistically significant sign on the coefficient estimates (betas). These betas are then exponentiated to get the multiplicative rate: anything above 1 indicates increase in infection rates, and those below 1 indicate a decrease in infection rates. We assume the following: no seasonality variations (daily time series data), a level shift effect (between days 5-10, rate of change of infection could be negative or positive but need to test statistical significance) for each intervention, and in some cases, the external effects of the interventions are accounted for.

## 3 Results and discussion

### 3.1 Estimating the true incidence rate by correcting for inconsistencies in testing rates

The Jack-knife method of estimating the incidence rate of COVID-19 infection has been applied to the data to correct for any inconsistencies in testing rates within the period under review (Overgaard et al., 2017; Rao et al., 2017). Ideally, to calculate the population (true) incidence rate, one needs to have consistent sampling rate (preferably per day) from a representative sample of the population, in which case the positivity rate would be expected to mirror the incidence rate. From Figure 2, the Jack-knife estimation is consistent with the actual COVID-19 case numbers observed in Ghana until May 7, 2020, from where the number of positives recorded is considerably higher than the Jack-knife estimates. This departure is consistent with the increased number of tests conducted on contact traced individual and concentrated within hotspots around that period, and neither general nor representative population level sampling. Thus, with increased testing targeted at hotspots and contacts of persons who tested positive, one would expect a higher number of positive cases. What this indicates is that there is a critical need to have consistency in testing rates, and sampling representative of the population, in order to generate a true incidence rate and furthermore, reproduction number. The forementioned estimates would form the basis of monitoring the impact of control interventions, which would inform critical policy decisions. In this regard, an exclusive focus on the actual number of positives after May 7 as the basis for measuring the state of affairs with regard to COVID-19 spread could be erroneous and lead to a false alarm.

**Figure 2:**
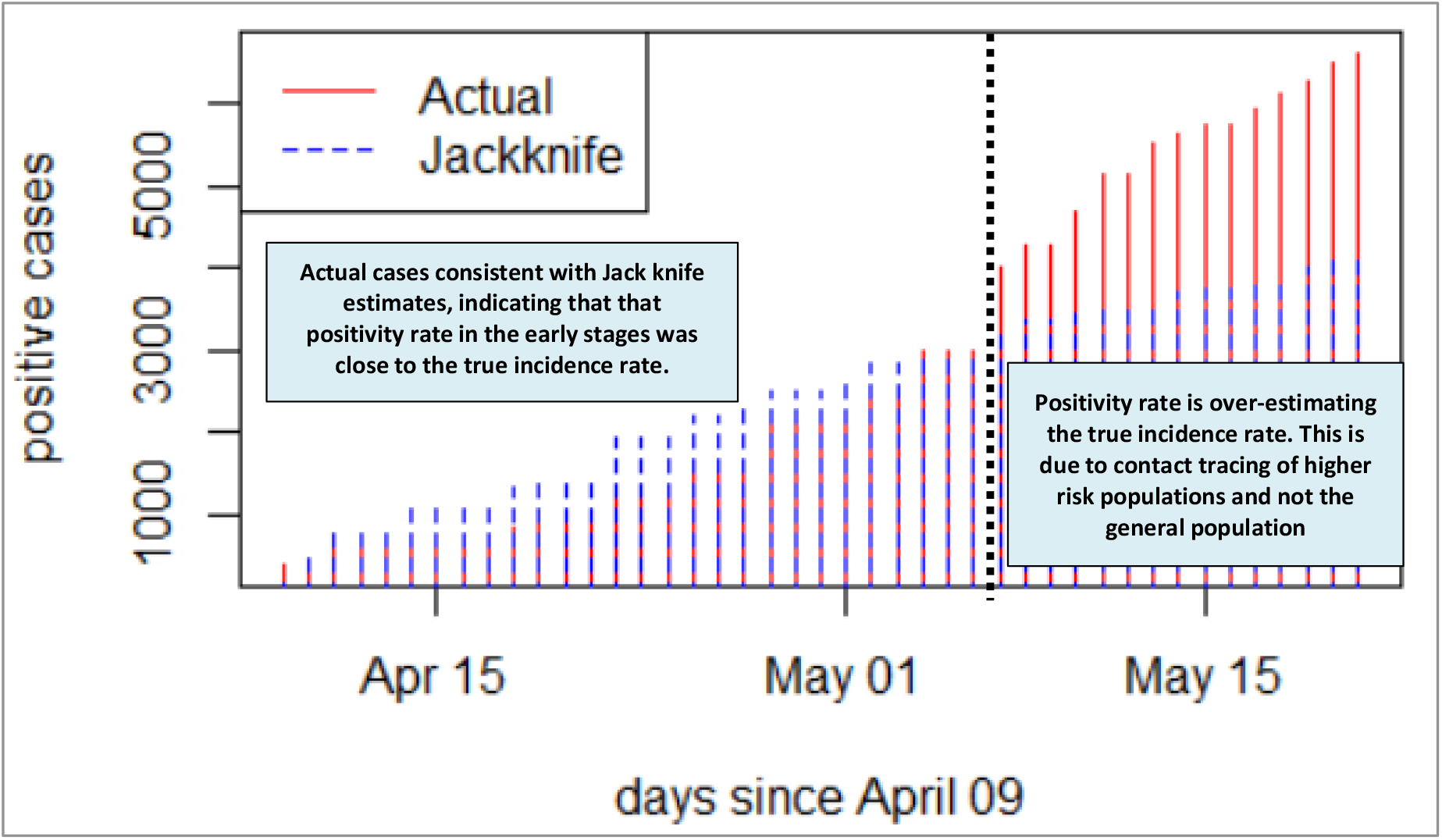
Jackknife estimates versus actual number of positive cases. Source: Authors’ estimates based on Ghana Health Service data

### 3.2 Initial reproduction number (*R*_*0*_)

Several growth models for the initial outbreak of COVID-19 in Ghana were considered and presented in Table 1.

**Table 1:** Estimates of different parameter models.

| Growth Model | Estimates (standard errors) | Akaike Information Criterion (AIC) |
| --- | --- | --- |
| Exponential | 0.095(0.002)** | -16914.29 |
| Weibull | 2.15(0.062)** | -16946.64 |
| Logistic | 0.17(0.006)** | -16991.13 |
| Gompertz | 0.147(0.0051)** | -17011.42 |

\*\* significant at 5%

The estimated models are shown below:

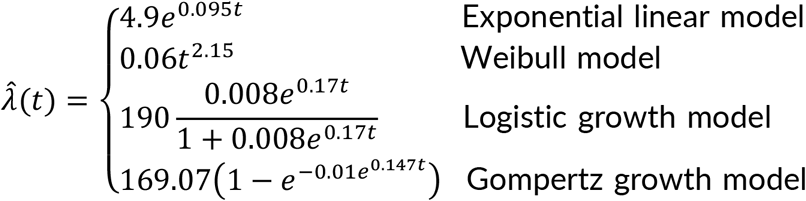

From Table 1, the underlying assumption behind the models is that in the initial stage of the epidemic, the number of infected individuals emerges at an exponential rate. We observe that the estimated growth rate for each model is statistically significant at 5% with the Gompertz model having the minimum AIC although all models were significantly close. The estimated growth rate from this model is 0.147 with 95% C.I. [0.137-0.157]. From Equation (8), assuming a 15-day time to recovery after COVID-19 infection^8^, the initial reproduction number is estimated as 3.205. That is: *R*_*0*_ = 1 + (0.147 * 15) = 3.21.

Table 2 shows our estimates of the initial reproduction number for Ghana compared with other countries. Ghana’s initial reproduction number is very close to that reported for South Africa and Morocco, and also relatively close to Kenya, Algeria but further afield to figures from some Chinese provinces and South Korea. This result is also note-worthy for one key reason: during the early days of the pandemic, Ghana was testing relatively larger numbers than several African countries at a good rate by adopting a pooled sampling testing approach, which allowed a lot more COVID-19 tests to be conducted (WHO, 2020b; Nyazika et al., 2020). Ghana, for example, is reported as having conducted over 370,000 tests between March and mid-July 2020, putting the country among the highest in Africa with regard to number of tests when standardised by the population size (WHO, 2020b). Generally speaking, countries testing at a good rate were getting R_0_ greater in the high 2s and low 3s.

**Table 2:** Estimates of R0 and comparison to other country studies.

| Country | Initial Reproduction Number ($R_0$) Estimate | Source |
| --- | --- | --- |
| Ghana | 3.21 | Authors' estimate |
| South Africa | 3.20 | Zhao et al. (2020) |
| Egypt | 2.30<br>2.29 | Desouky (2020)<br>Zhao et al. (2020) |
| Algeria | 2.66 | Zhao et al. (2020) |
| Morocco | 2.99 | Hammoumi & Qesmi (2020) |
| Kenya | 2.82 | Zhao et al. (2020) |
| Nigeria | 2.29 | Zhao et al. (2020) |
| China | 4.03 (Hubei Province)<br>3.80 (Wuhan)<br>2.60 (Wuhan)<br>2.68 (Wuhan)<br>2.20 (Wuhan)<br>2.61 (China) | Choi & Ki (2020)<br>Read et al. (2020)<br>Anastassopoulou et al. (2020)<br>Wu et al. (2020)<br>Li et al. (2020)<br>Wang et al. (2020) |
| South Korea | 3.47-3.54 (Daegu/NGP) | Choi & Ki (2020) |
| United Kingdom | 2.6 | Jarvis et al. (2020) |

### 3.3 Time dependent and Sequential Bayesian reproduction number

It will be necessary for the generation time distribution to be discretised using the same time unit. We therefore assumed a Gamma distribution for the generation time distribution with mean 3.57 and standard deviation 2.55 (Davies et al, 2020a; Davies et al., 2020b) when there are control measures in place. We also assumed that the infectious period is the same for asymptomatic and symptomatic cases. These methods require a time period over which there is an exponential growth.

The time period chosen is the date from first case(s) (March 12, 2020; day 1) to the date of maximum incidence (April 25, 2020; day 44) (Figure 3). From the two outputs in Figure 3, the Sequential Bayesian does not make biological reasoning because the assumption of equal probabilities of *R*_*0*_ does not hold (Clancy & O’Neill, 2008). Hence, we opt for the time-dependent method.

**Figure 3:**
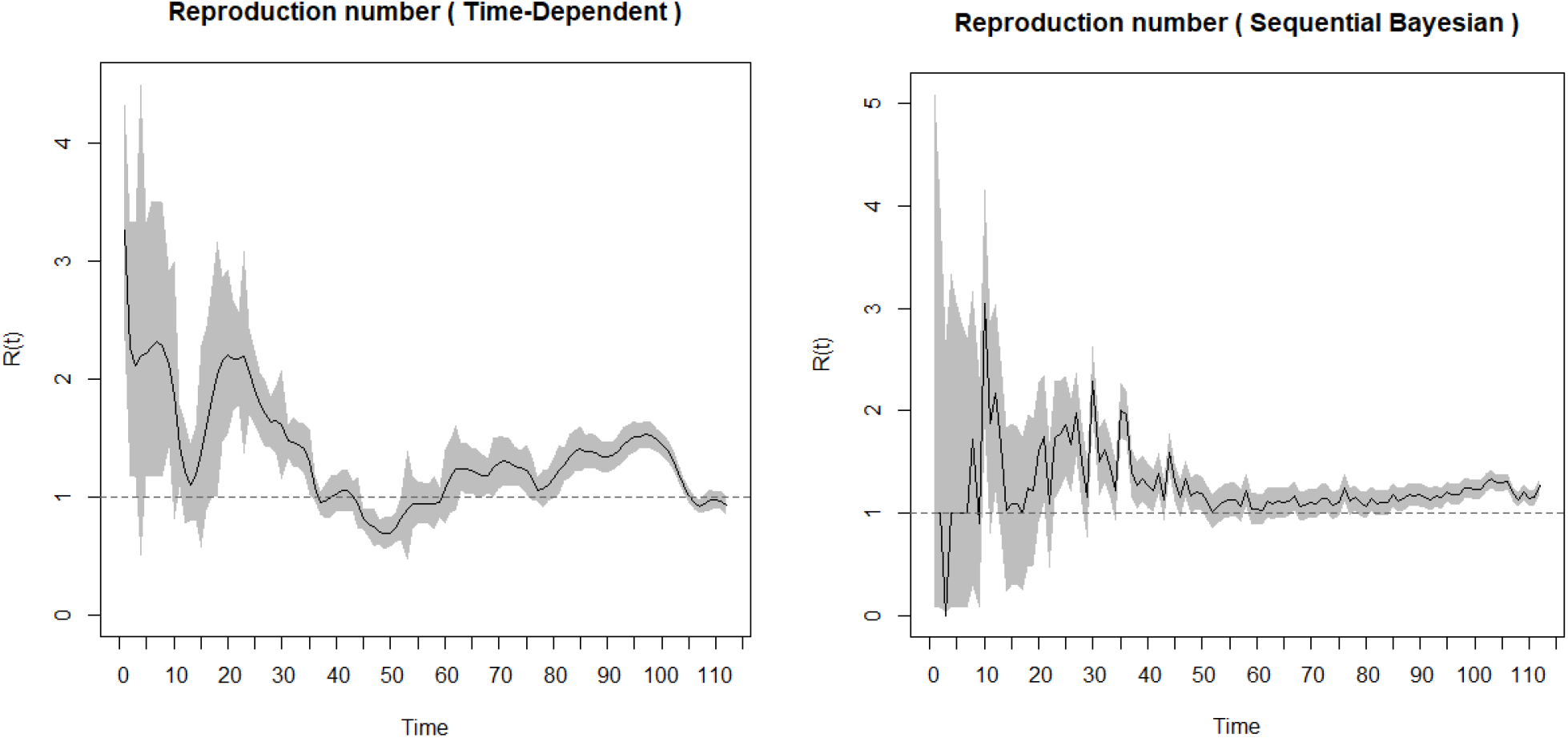
(a)Time dependent and (b) Sequential Bayesian reproduction number. Source: Authors estimates, based on Ghana Health Service data

From Figure 4, the first estimates of *R*_*0*_ in Ghana reflect the high rate of spread at the onset (March 12, 2020) of the pandemic outbreak. This was right before the mandatory quarantine of all persons entering the country, including events such as the ban on public gatherings, closure of schools, churches, mosques, and restriction of entry into Ghana for anyone who had been in a country with more than 200 COVID-19 cases. The spread of the disease measured by the *R*_*0*_ then begins to decline after those key events from the first reported date (March 12, 2020) to the date of border closure (10 days interval). Between the date of border closure and right before the announcement of the partial lockdown of two regions (greater Accra, greater Kumasi), which reported more than 90% of the cases, the spread of the disease increased moderately. After the initial lockdown, the spread of the disease reduced initially but begun to pick up momentum again from mid-April onwards with the easing of restrictions. This was despite the introduction of the wearing of mandatory facemasks, which were not fully complied with by citizens due the largely informal nature of settlements in the country making enforcement even more challenging.

**Figure 4:**
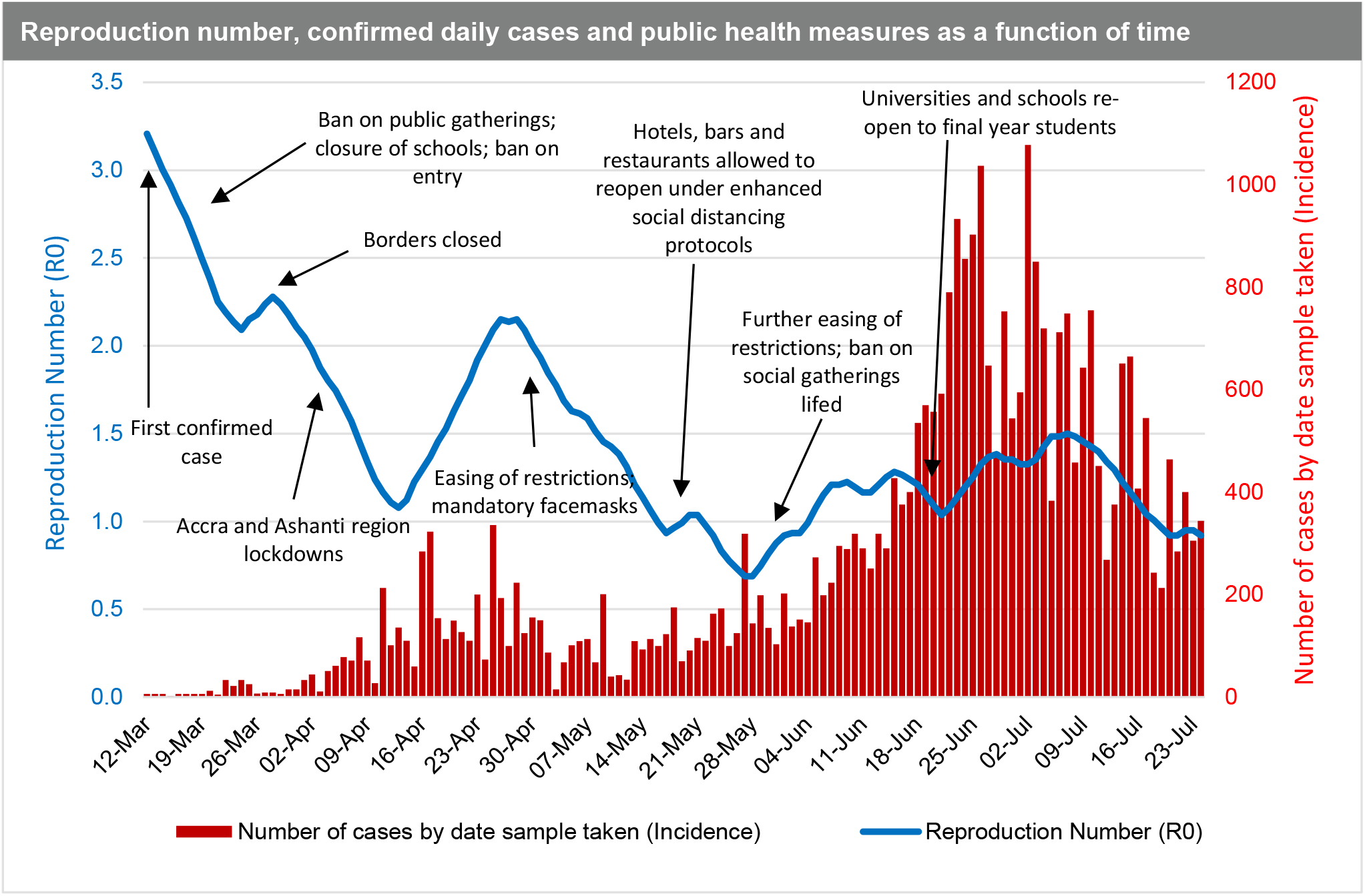
Time dependent R_0_ superimposed on daily COVID-19 infections in Ghana. Source: Authors estimates, based on Ghana Health Service data

In essence, the reproduction number reduces from March until mid-April during the lockdown, and then upsurged again when the country relaxed the initial restrictions. However, we note that the *R*_*0*_ decreases further even after restrictions are eased (including when bars and hospitality venues were allowed to operate). With this observation, it can be inferred that the reproduction number and incidence rate paint an accurate picture of the COVID-19 spread in Ghana at the early stages of the pandemic from March to early May 2020. In these early stages, Ghana’s actual cases of COVID-19 are consistent with the Jack-knife estimates (see Figure 2), indicating that that the positivity rate in the early stages was close to the true incidence rate. Beyond the early stages of the pandemic, it becomes difficult to make any deductive inferences based on the data available which might reflect the true situation. Our argument is further supported by that fact that contrasting to the *R*_*0*_, the positivity rate, which ideally should be a proxy for the incidence rate, had been on the ascendency since mid-April (Figure 5).

**Figure 5:**
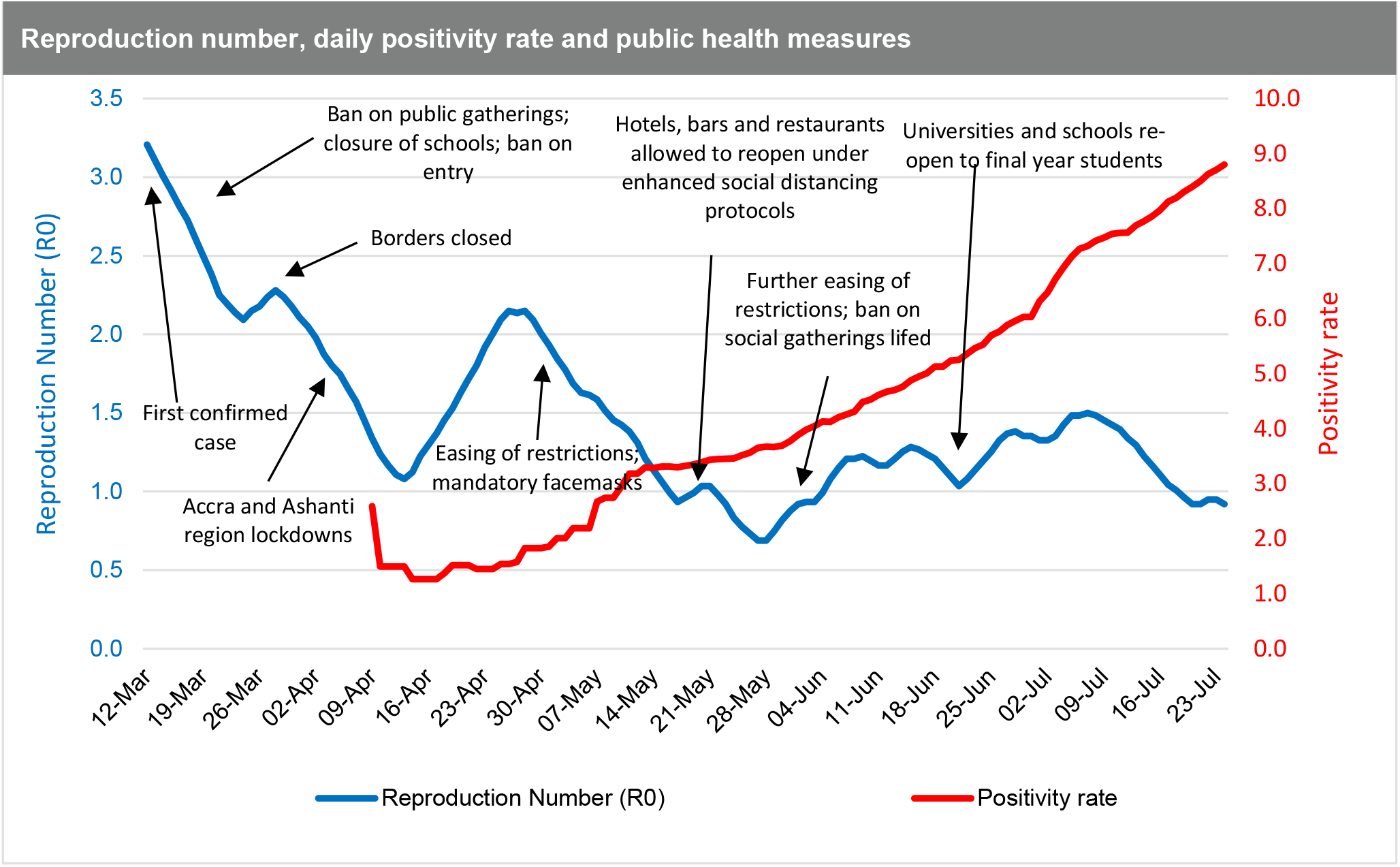
Time dependent R_0_ superimposed on daily positivity rate in Ghana. Source: Authors estimates, based on Ghana Health Service data

In essence, the reproduction number estimated will be unreliable if a country does not have a consistent and population representative testing regime. In Ghana’s case, the reproduction number exhibits significant volatility as can be seen in the time-dependent charts (Figures 4 and 5), indicating that it is not representative of the true progression of the epidemic, as there was not a consistent testing regime in place. The reproduction number is a function of the frequency of potential exposures, and not having a consistent testing regime in place means that any such estimate is subject to misrepresentation and will be ineffective for use in evidence-based policy making. In fact, a very high positivity rate might actually be an indication that a country is not doing enough COVID-19 testing within the general population aimed at finding and isolating as many cases as possible in order to curb transmission.

### 3.4 Impact of easing restrictions on COVID-19 infection rates

Next, we look at the intervention periods (Figure 6) and their mitigation effect on the rate of infections. As discussed in **Section 2.2.3**, we fitted a Negative-Binomial model with the log link function, by taking into account serial dependence which includes a regression on the two successive previous observations, and also include the six intervention effects detected by the model (Fokianos, 2015). Using this approach, we are able to assess the effect of interventions such as easing restrictions, on COVID-19 infection rates and to separate these from general trends and serial dependencies in time (Gilmour et al.,2006).

**Figure 6:**
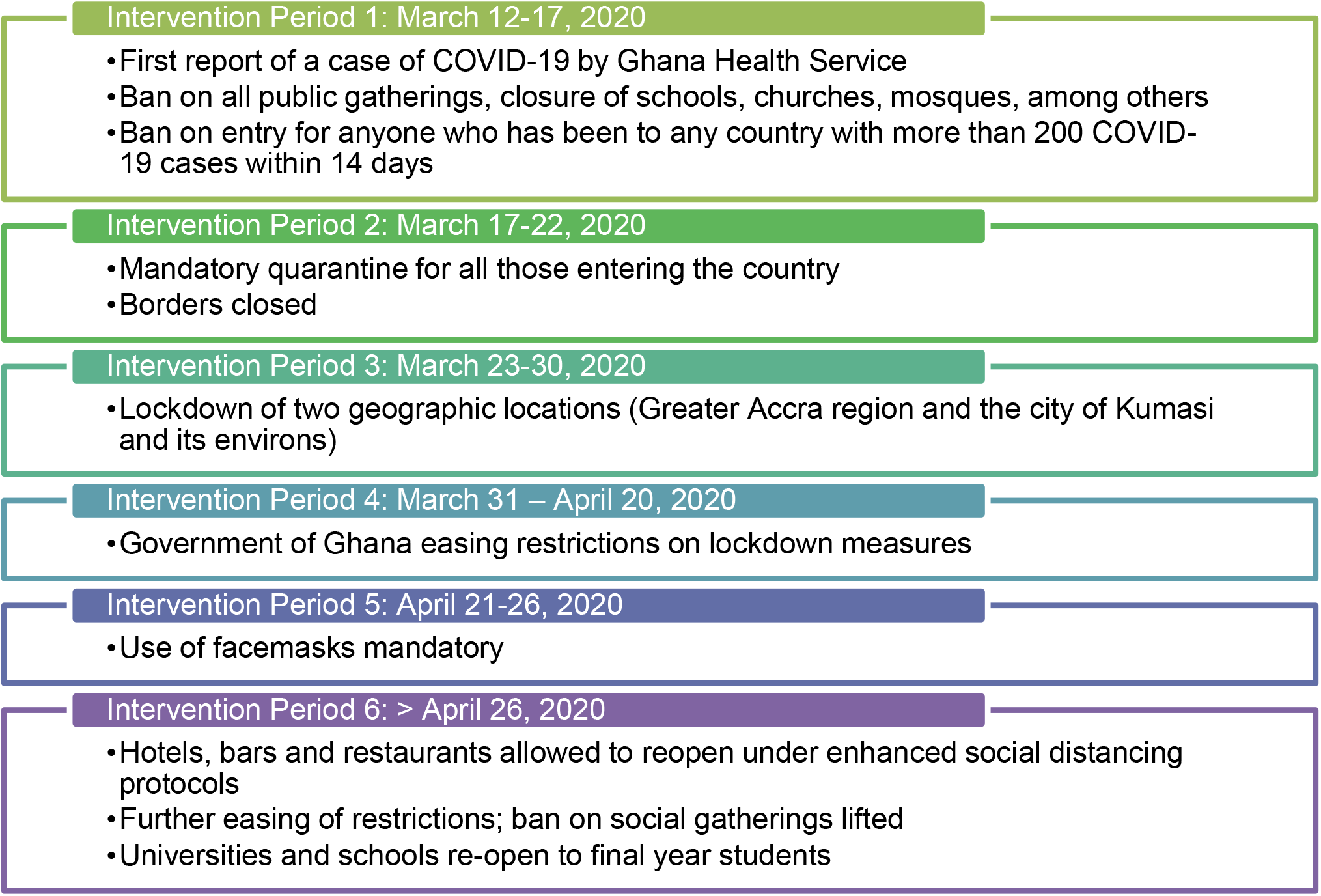
Summary of intervention periods and public health measures imposed. Source: Authors’ depiction

As can be seen in Table 3, there is a mixed a story of the impact of the mitigation effects of easing restrictions on COVID-19 infection rates. The initial three weeks following the confirmation of the first case on March 12 was characterised by a 25% increase in the rate of infection although this not statistically significant (Intervention Period 1). During this time, the Ghana government swiftly moved to impose a ban on all public gatherings, closure of schools, churches, mosques, among others. It also banned entry for anyone who has been to any country with more than 200 COVID-19 cases within 14 days. Intervention Period 2 was accompanied by the policy announcement of the mandatory quarantine for all those entering the country and the closure of air and land borders. The period was characterised by a 6% decrease in the rate of infection, but this was again not statistically significant impact.

**Table 3:** Testing for intervention effects. The null hypothesis (H_o_) is no intervention effect.

| Date | Intervention Type | Time(days) | Coeff. Estimates | Multiplicative effect (rate of infection) | Comment<br>Below 1 = decrease<br>Above 1 = increase |
| --- | --- | --- | --- | --- | --- |
| March 12-17 | Intervention Period 1 | 5<intervention≤10 | **0.223** | Exp (0.223) = 1.250 | 25% increase in the rate of infection. However, this not statistically significant impact |
| March 17-22 | Intervention Period 2 | 10<intervention≤18 | **-0.066** | Exp (-0.066) = 0.936 | 6% decrease in the rate of infection. However, this not statistically significant impact |
| March 23-30 | Intervention Period 3 | 18<intervention≤38 | **0.445** | Exp (0.445) = 1.560 | 56% increase in the rate of infection. However, this not statistically significant impact |
| March 31 – April 20 | Intervention Period 4 | 38<intervention≤44 | **-0.382\*\*\*** | Exp (-0.382) = 0.682 | Statistically significant 32% decline in rate of infection. |
| April 21-26 | Intervention Period 5 | 44<intervention≤79 | **0.129** | Exp (0.129) = 1.138 | 14% increase in rate of infection. However, this not statistically significant impact |
| >26 April | Intervention Period 6 | Intervention>79 | **0.043** | Exp (-0.025) = 1.044 | 4% increase in rate of infection. However, this not statistically significant impact |

Source: Authors' estimates | \*\*\* highly significant (<0.001)

Following this, Intervention Period 3 saw the government move to further impose the lockdown of Ghana’s main population centres: the two geographic locations — Greater Accra region (5.06 million) and the city of Kumasi and its environs (3.4 million residents) — which collectively account for about 27% of the country’s 31 million population as of 2020. At the time of Intervention Period 3, we also see a 56% decrease in the rate of infection. However, this not statistically a significant impact. The Ghana government begun easing restrictions on these earlier lockdown measures imposed by Intervention Period 4. However, we observe a statistically significant 32% decline in rate of infection relative to previous intervention period. We explain this trend as possibly due to the residual positive effect of the preceding intervention period now kicking in, or due to a lag in the actual resumption of mass gatherings, as all had been cancelled and were either yet to be rescheduled or simply did not come off any longer. These are also confirmed in several studies, notably Meiksin (2020), Li et al. (2021) and Shengjie et al. (2020). For example, Meiksin (2020) notes in the context of the United Kingdom that the weekly death rate of registered COVID-19 deaths peaked just over three weeks after the partial lockdown and continued to decline. Li et al (2021) also found reduced COVID-19 transmission following the introduction of non-pharmaceutical interventions such as school closure, workplace closure and public events ban of between 3% to 24% on day 28 (four weeks) following the introduction, compared with the last day before introduction. Likewise, Shengjie et al. (2020) found that non-pharmaceutical interventions which were deployed in China were effective in containing the COVID-19 outbreak although the efficacy of the different interventions was dependent on early case detection and effective contact reduction.

Intervention Periods 5 and 6 were also occasioned by increases in the rate of infection as many citizens begun to fully get back to their normal activities and despite the use of facemasks now made mandatory. For example, hotels, bars and restaurants being allowed to reopen under enhanced social distancing protocols as part of further easing of restrictions. Additionally, the ban on social gatherings was lifted and universities and schools were re-open to final year students.

## 4 Conclusions

The data available suggests that the COVID-19 lockdown and other restrictions in Ghana did actually reduce movement and interaction which would in turn had a positive effect in slowing down the spread of the virus in the early phase of the pandemic. This conclusion is further supported by anonymised and aggregated data from Mobile Network Operators on the mobility patterns of populations published by the Ghana Statistical Service for the period between February 17 and May 03, 2020 showing that person movements decreased by up to 60% relative to the baseline value during the period when restrictions were in place.

The initial reproduction number (*R*_*0*_) is estimated from the statistical model as 3.21 using a 0.147 estimated growth rate and a 15-day time to recovery after COVID-19 infection. We observe that the estimated growth rate for each model is significant at 5%, with the Gompertz model having the minimum AIC, although all models were significantly close. The estimate of the initial *R*_*0*_ is consistent with what is reported in the literature from other parts of Africa, China and Europe. The *R*_*0*_ reflects the high rate of spread at the outset of the pandemic in the country before the mandatory quarantine of all persons entering the country as well as the initial ban on public gatherings, among other public health interventions. The *R*_*0*_ estimates and incidence rates could be seen as a fair reflection of the COVID-19 epidemiology at the early stages of the pandemic in Ghana (from March to early May 2020). From our analysis, Ghana’s actual cases reported are consistent with the bootstrap Jack-knife estimates, indicating that that positivity rate in the early stages closely mirrored the incidence rate.

Going forward, there is a need more robust, consistent, systematic and representative sampling for testing, first to enable early detection, isolation and treatment of infected individuals and to reduce progression to severe disease and risk of mortality. Second, our recommended testing regime is critical for generating data for monitoring the impact of interventions to inform policy decisions, especially in this period of looming austerity.. We therefore recommend the widespread deployment of point-of-care diagnostics and later rapid diagnostic tests (RDT) – when they become more reliable, coupled with timely reporting and dissemination of results, as a first-line strategy towards mass surveillance of the population both to monitor the reproduction number and estimate the true incidence rate of COVID-19. The fore-mentioned measures are not only logistically less burdensome but would also prove timelier and more cost effective than running full-sample or pooled PCR tests.

Monitoring the impact of control measures on the evolution of the pandemic in Ghana will require being able to determine any changes in transmission rates, hospitalization rates and death rates over time. Each of these measures provide unique insights for disease control and prevention policy making and implementation but can only be determined by consistent, systematic and population representative testing mechanisms. Authorities would therefore need to rapidly set up a nationally representative monitoring systems involving sentinel sites, including schools, markets, places of worship, mass transit stations and other places of gathering in each of the 16 regions of Ghana. From these sentinel sites, a regular number of tests via an algorithm will need to be conducted (preferably on a daily basis), with results made available within 24-48hrs. Results from this system could serve as the index for measuring the infectivity rate and subsequently, an accurate-enough time-dependent *R*_*0*_ on a daily and weekly basis in different parts of the country.

While the results presented in this paper provide important insights into the early evolution of the pandemic in Ghana, we note that Ghana’s daily incidence data at the early stages which has been used in the analysis, was sparse, lacked granularity and did not show a consistent trend. This means our assumptions of the disease parameter estimates must be applied cautiously to policy making, while improvements in sampling and testing strategies are improve as we have recommended.

## Data Availability

The data used in this study and R modelling code can be obtained on written request to the corresponding author.

## Authorship statement

All authors certify that they meet the criteria for authorship of the ICMJE

## Funding

This research received no external funding.

## Declarations of Competing Interests

Authors have no conflicts of interest to declare

## Footnotes

8 CDC (2020). Interim Guidance on Duration of Isolation and Precautions for Adults with COVID-19Available at: https://www.cdc.gov/coronavirus/2019-ncov/hcp/duration-isolation.html (Accessed: 10 February 2021).

## Notes

### Competing Interest Statement

The authors have declared no competing interest.

### Author Declarations

Kumasi Center for Collaborative Research in Tropical Medicine (KCCR), Kumasi, Ghana.

